# The spatio-temporal epidemic dynamics of COVID-19 outbreak in Africa

**DOI:** 10.1101/2020.04.21.20074435

**Authors:** Ezra Gayawan, Olawale Awe, Bamidele M Oseni, Ikemefuna C. Uzochukwu, Adeshina Adekunle, Gbemisola Samuel, Damon P Eisen, Oyelola A Adegboye

## Abstract

The novel coronavirus (COVID-19) caused by severe acute respiratory syndrome coronavirus 2 (SARS-CoV-2), emerged in the city of Wuhan, China in December 2019. Although, the disease appears on the African continent late, it has spread to virtually all the countries. We provide early spatio-temporal dynamics of COVID-19 within the first 62 days of the disease’s appearance on the African continent. We used a two-parameter hurdle Poisson model to simultaneously analyze the zero counts and the frequency of occurrence. We investigate the effects of important healthcare capacities including hospital beds and number of medical doctors in the different countries. The results show that cases of the pandemic vary geographically across Africa with notable high incidence in neighboring countries particularly in West and North Africa. The burden of the disease (per 100,000) was most felt in Djibouti Tunisia, Morocco and Algeria. Temporally, during the first 4 weeks, the burden was highest in Senegal, Egypt and Mauritania, but by mid-April it shifted to Somalia, Chad, Guinea, Tanzania, Gabon, Sudan, and Zimbabwe. Currently, Namibia, Angola, South Sudan, Burundi and Uganda have the least burden. The findings could be useful in implementing epidemiological intervention and allocation of scarce resources based on heterogeneity of the disease patterns.

## Introduction

On March 11, 2020, the World Health Organization (WHO) declared the novel coronavirus disease (COVID-19) outbreak a pandemic. The disease, caused by severe acute respiratory syndrome coronavirus 2 (SARS-CoV-2), emerged in the city of Wuhan, China in late December 2019 and has quickly spread globally with 1,436,198 cases and a case fatality rate (CFR) of 5.96% as of April 9, 2020. [1]. This pandemic has not only become a public health crisis leading to loss of life but has caused the global economy to melt down with severe disruption to international travel, tourism and trade. As of April 7, 2020, 52 African countries have reported 10,086 confirmed cases and 487 deaths from the pandemic yielding a CFR 3.38%. It is likely that the case ascertainment in Africa is incomplete.

Human population movement generally plays an important role in the spread of infectious diseases and this particularly applies to COVID-19 as this respiratory virus is highly transmissible. The reasons for the late appearance of COVID-19 in Africa compared with other parts of the world are unknown but it may be due to relatively limited international travel to the continent [2]. There has always been awareness of importation of infectious diseases such as Ebola and tuberculosis from Africa to the West [3, 4]. Largely, African countries have reported their first COVID-19 cases to be imported from European countries, possible due to the proximity of the two continents [5]. The first case was confirmed on the African continent on February 14th, 2020 in Egypt followed by Nigeria on February 27th. The initial dynamics of the disease demonstrated slow spread across the continent until the situation escalated quickly in the last week of March.

Now, during the COVID-19 pandemic, global experts have shown concern about the spread of this disease in Africa, because of grossly underfunded and inadequate healthcare systems. Early detection and control of outbreaks is inefficient and unreliable due to poor disease surveillance, insufficient training of healthcare workers, and inadequate data transmission [2, 6-8].

Within the frame of the outbreak of the COVID-19 pandemic, there have been a number of applications of statistical models to predict infection rate and spread [9, 10]. However, mapping of disease incidence to identify spatial clustering and patterns remains an important pathway to understanding disease epidemiology and is required for effective planning, prevention or containment actions [11-13]. There are a few studies that attempt to map the pandemic in China [14] and in Iran [15]. However, the temporal dynamic of the pandemic has not been taken into account in order to access the space-time dynamics. Further, to our knowledge, there is dearth of studies that have examined the spatio-temporal dynamics in Africa. Our aim, therefore, is to analyze the spatio-temporal dynamics of COVID-19 within the first 62 days of the disease’s arrival on the African continent. We propose a two-parameter hurdle Poisson model to simultaneously analyse the zero counts as well as average occurrence of the disease. The two parameters are extended, through appropriately chosen link functions, to the spatio-temporal covariates following the framework of distributional regression coined by Klein et al. [16]. Additionally, we investigate the effect of important healthcare capacities including hospital beds and the number of medical doctors on the risk of COVID-19 in the different African countries. The hurdle model is a modified count model in which two processes generating the zeros and the positives are not constrained to be the same [17]. The idea is that a binomial distribution model governs the binary outcome that stipulates whether the count variable returns a zero or a positive realization. For the truncated-at-zero count data, a conditional distribution, in this case, a Poisson distribution is considered. With this, we are able to examine both the patterns of zeros and the average counts of the pandemic across space and time throughout Africa.

## Methods

### Data sources

In this paper, we used publicly available daily number of confirmed COVID-19 cases reported by the World Health Organization (https://covid19.who.int) from 14th February to 15th April 2020. Due to the requirement of the spatial effect model considered in this study, we only included 47 African countries that have confirmed COVID-19 cases and share at least one international boundary with another country. Additionally, we obtained data on healthcare capacities: number of hospital beds and physicians for each of the countries from the World Development Indicators of the World Bank (https://data.worldbank.org).

### Statistical analysis

Preliminary exploratory spatial analysis was used to investigate the spatial and spatio-temporal distribution of incidence of COVID-19, healthcare capacities (number of hospital beds and number of physicians) across Africa. We used Pearson’s correlation to assess the relationships between number of confirmed COVID-19 cases and each of the two healthcare capabilities of each country.

For the spatio-temporal analysis, we considered a two-component hurdle Poisson model which consists of a point mass at zero followed by a truncated Poisson distribution for the non-zero count observation. For an independently and identically distributed random variable, the hurdle Poisson distribution is expressed as

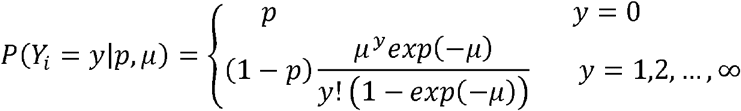

where *Y*_*i*_ is the response variable of interest that counts the occurrence of COVID-19 cases at a particular country, is the none occurrence probability, that is, the probability of not reporting a COVID-19 case in a given, day and *μ* measures the frequency of occurrence. Thus, as increase, the average count of COVID-19 increases. If *p* is 0, it implies that each country reported an infection during the period under consideration, but if is 1 then no one would be infected by the pandemic on the continent. Usually, *p* is considered to be strictly between 0 and 1, such that everyone in the population of the African continent has a non-zero probability of being infected with the virus even if they do not get infected during the period considered. Under the hurdle distribution, the expected value of *Y* is given by *E* (*Y*) = *pμ*/(1 – *exp*(−*μ*)).

Based on the framework of distributional regression that allows the multiple parameters of a response distribution, rather than just the mean as common in most classical applications, we extend the two parameters space ϑ_*k*_ =(ϑ_1_=*p*,ϑ_2_ = *μ*) of the hurdle Poisson model to a the spatial and spatio-temporal covariates of the COVID-19 cases in Africa through some suitable (one-to-one) link functions that ensure appropriate restrictions on the parameter space.

The general form of the geo-additive hurdle Poisson model considered is given by

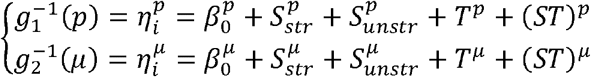

where *g*_1_ and *g*_2_ are link functions chosen as logit and log links for the parameters *p* and *μ* respectively. Omitting subscript, *β*_0_ is the model constant term, *S*_*str*_ is the structured spatial random effect, *S*_*unstr*_ is the unstructured random effect, *T* is the temporal term and *ST* accounts for the spatio-temporal random effect. Consequently, the temporal term was model based on Bayesian P-spline. This allows for the estimation of the temporal term as a linear combination of basis spline (B-splines) based on a second order random walk prior with inverse gamma for the hyperparameters [18]. We considered cubic B-splines with 20 equidistance knots, which induce enough flexibility to capture even the most severe non-linearity.

For the all structured spatial and spatio-temporal effects, the countries are considered as discrete set of spatial locations and we used a Markov random field prior that considers a binary structure for the neighborhood structure of the countries such that proximate locations that share boundaries are assigned a weight of 1 and 0 if they do not. To ensure smoothness, we consider a Gaussian Markov random field prior that induces a penalty in which differences between spatially adjacent regions are penalized. Exchangeable independent and identically distributed normal prior was considered for the unstructured random effects.

The Bayesian inference is based on the distributional regression framework of Klein, Kneib [16], who developed some Markov chain Monte Carlo simulation techniques in which suitable proposal densities are constructed based on iterative weighted least squares approximation to the full conditional. All smoothing variance parameters and hyperparameters are assigned inverse gamma hyperpriors and we perform some sensitivity analysis but the results, based on the different hyperpriors, turn out to be indistinguishable.

To implement the spatio-temporal component, we considered weekly distributions of the pandemic across the continent but due to paucity of data during the first weeks of occurrence, we considered the first month (14th Februsary-13th March) as the first week. Thus, the spatio-temporal complete data were grouped into a six-week period. The intention was to examine how the countries fared in terms of the occurrence of the pandemic over a weekly period. We implement four models, by sub-setting temporal and spatial covariates on the mean parameter while keeping the temporal, structured and unstructured spatial effects for the probability parameter, and based model choice on deviance information criterion (DIC). The details of the implemented models including the values of the DIC are presented in Table 1. For all models, we performed 15,000 iterations with 3,000 set as burn-in and the thinning parameter was set at 10. The generated Markov chains were investigated through trace plots to ascertain mixing and convergence.

**Table 1.** Assessment of various model specification used in this study^†^

| Model | Specification | $\bar{D}$ | pD | DIC |
| --- | --- | --- | --- | --- |
| 1 | Spatial ($S_{str} + S_{unstr}$) + ST | 12013.66 | 219.49 | 12452.66 |
| 2 | Trend + ST | 9661.47 | 317.98 | 10297.43 |
| 3 | Trend + spatial ($S_{str} + S_{unstr}$) | 12237.92 | 107.30 | 12452.53 |
| 4 | Trend + spatial ($S_{str} + S_{unstr}$) + ST | 9134.13 | 215.38 | 9564.89 |

<sup>†</sup> $S_{str}$ represents structured spatial random effect, $S_{unstr}$ is the unstructured random effect, *Trend* is the temporal term and *ST* accounts for the spatio-temporal random effect.

## Results

### Preliminary COVID-19 distribution in Africa

The distribution of the present state of COVID-19 as at 11th April, the number of hospital beds and physicians (per 10,000 population) by country are presented in Figure 1(A-C). The Figure shows that cases of COVID-10 varied geographically across Africa with notable high incidence in West and North Africa (Figure 1A). However, when this incidence was converted to number of cases per 100,000 population, the burden of the disease in Africa was most felt in Djibouti, East Africa and North Africa (Tunisia, Morocco and Algeria) (Figure S1). Interestingly, countries with the highest burden of the pandemic in Africa are among those with the highest number of hospital beds and physicians, particularly those from the northern fringe (Figure 1B-C). Figure 2 examines the pattern of relationships between the pandemic and numbers of hospital beds and physicians. Findings reveal a positive correlation between COVID-19 and each of number of physicians (r=0.49, p-value=<0.001) and hospital beds (r=0.14, p-value=0.34) though only the estimate for physicians is significant.

**Figure 1.**
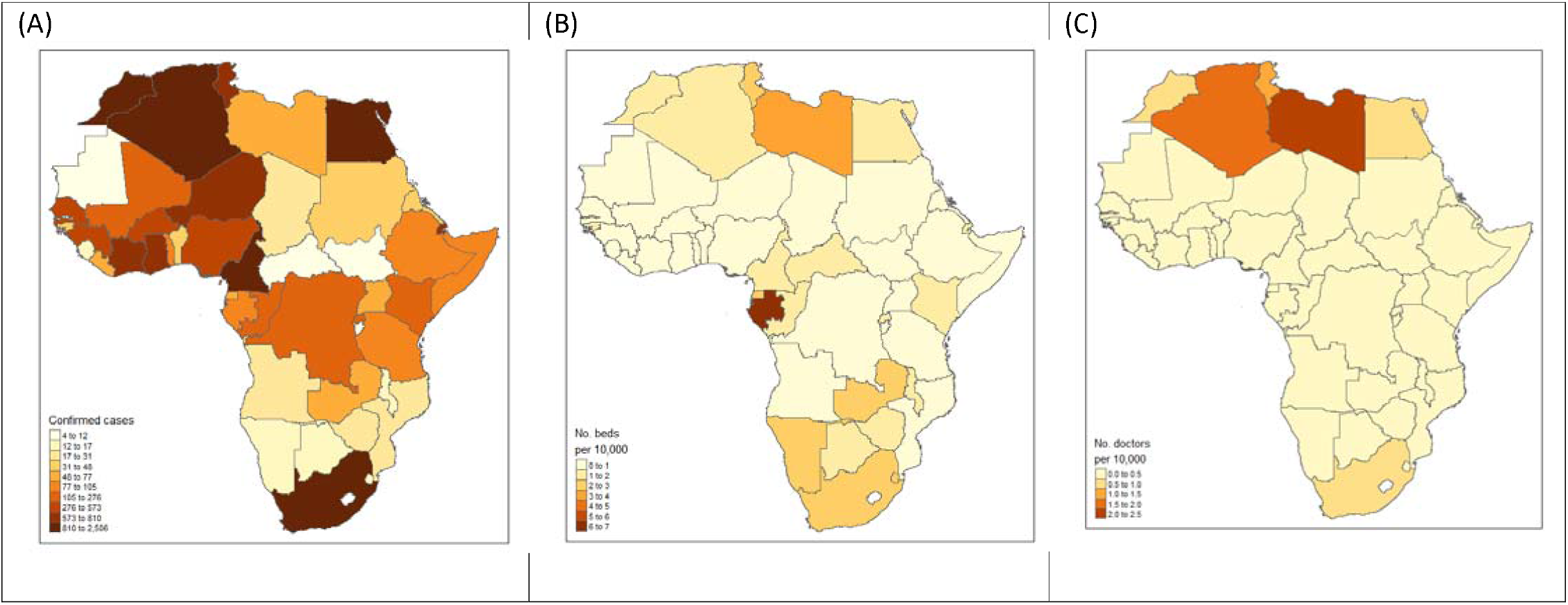
(A) Total number of confirmed COVID-19 cases as at 11^th^ April 2020, (B) Distribution of the number of hospital beds (per 10,000), (C) Distribution of the number of physicians (per 10,000).

**Figure 2:**
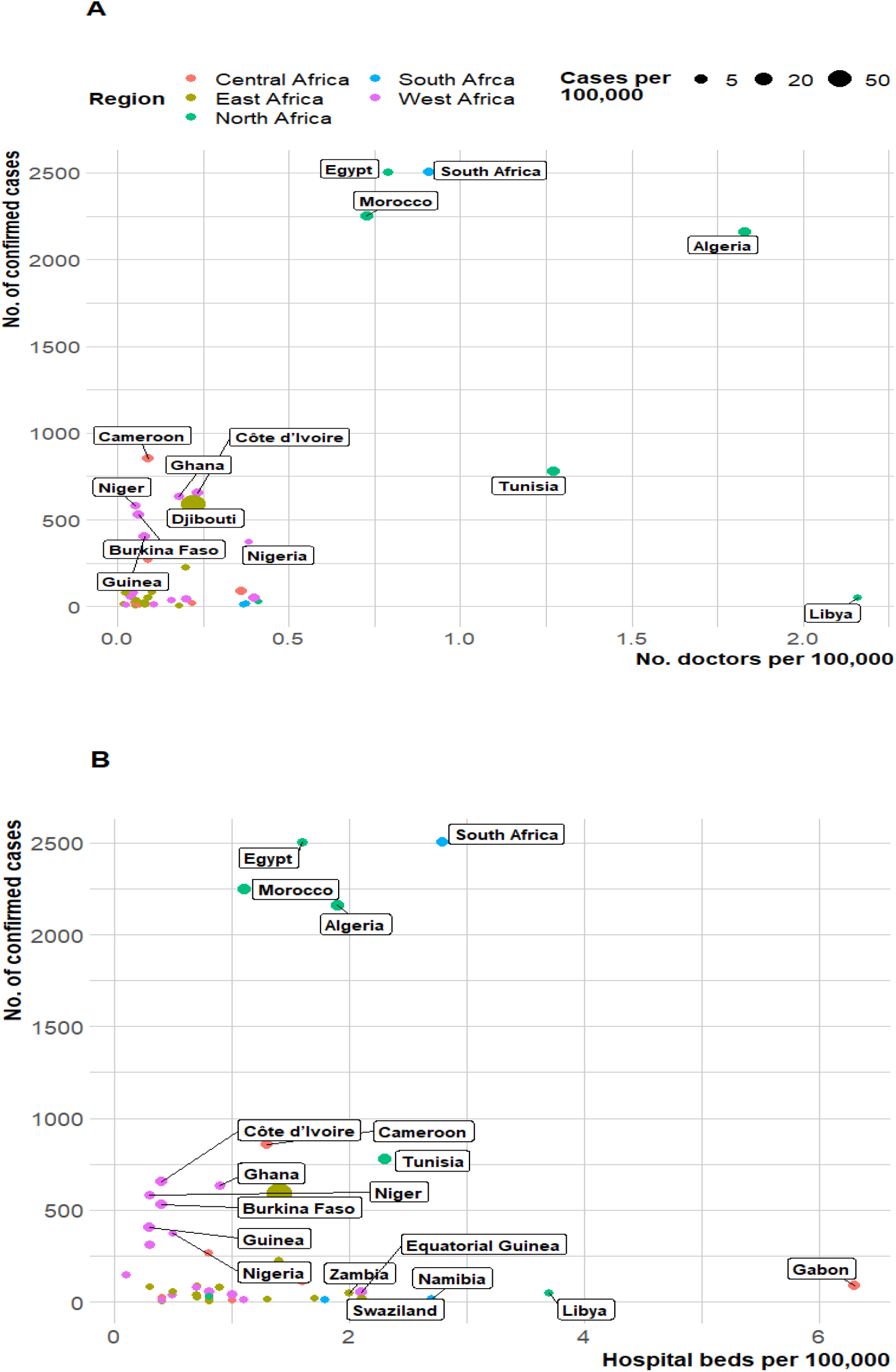
Scatter of number of confirmed cases of COVID-19 and healthcare capacities (Number of hospital beds/medical doctors).

### Spatio-temporal analysis

Table 1 presents the specifications of the four models considered together with the values of the model diagnostics criteria. As evident from the Table, the fourth model whose mean component includes the trend, structured and unstructured random effects, and the spatio-temporal components had the lowest DIC value and thus provides the best fit. Presentations of results shall therefore be based on those of this model. Figure 3 presents the maps of Africa showing the spatio-temporal patterns of the parameter *λ*, measuring the frequency of occurrence of COVID-19 on the continent during the period 14th February to 15th April, 2020, based on a six-week group.. The results show that during the period 14th February – 13th March, Senegal has the highest average record of the pandemic closely followed by countries such as Egypt and Mauritania. By the week of 14th -20th March, the burden shifted to Togo, South Africa, Egypt, DR Congo, Senegal and Burkina Faso. However, during the period 21st to 27th March, the findings show that South Africa had the highest burden of the pandemic, while for the week 28th March to, 3rd April, the share pandemic appears to be relatively similar across the countries. For the week 4th – 10th April, the burden shifted to Niger, Morocco, Guinea, Egypt, and Sierra Leone and lastly, for the week 11th – 15th April, the burden was most felt by countries such as Somalia, Chad, Guinea, Tanzania, Gabon, Sudan, and Zimbabwe but least for Namibia, Angola, and Uganda. The specifications and diagnostic criterions are shown in Table 1.

**Figure 3:**
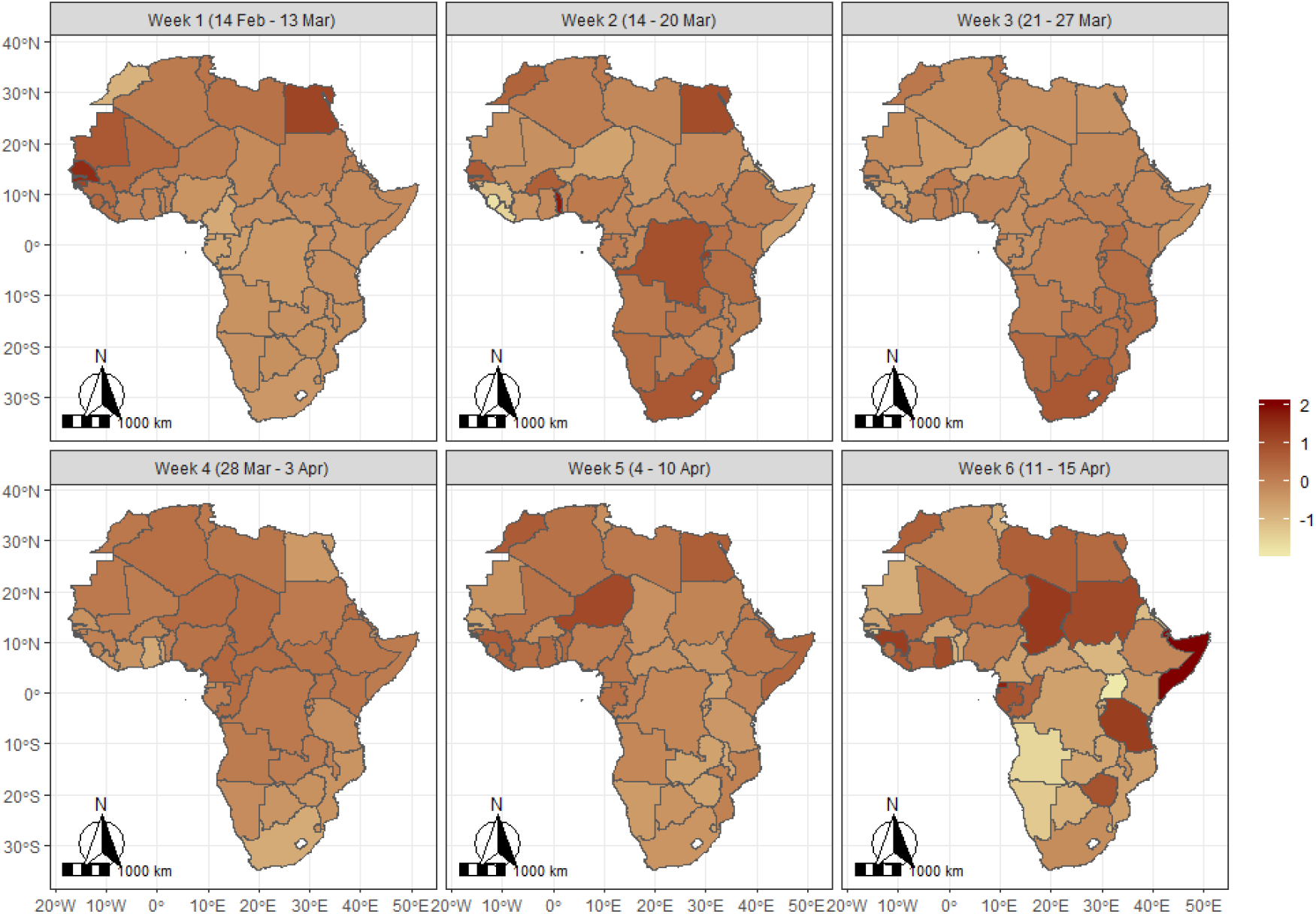
Spatiotemporal pattern of COVID-19 in Africa.

Results for the structured and unstructured random terms are presented in Figure 4 (A & B) for the frequency of occurrence of the pandemic. The two maps reveal different patterns across the continent, obviously because neighborhood structure of the countries was taken into account only in the structured effect. Thus, the structured random effect presents a western-southern divide indicating that the burden of the pandemic has been generally heavier among countries in the West African region specifically, in neighboring Ivory Coast, Burkina Faso, Ghana, Mali, Guinea, Senegal, as well as Morocco and Algeria in North Africa but generally lighter in the southern African countries. However, estimates from the unstructured effect that assumes independent and identically distributed normal prior show that South Africa, Egypt, Algeria, Morocco, Tunisia, and Cameroon, had the highest individual burden but lowest for South Sudan, Central African Republic, and Mauritania. The estimates are moderate for Nigeria, Ghana, Ivory Coast, Burkina Faso, Niger, Senegal, Republic of Congo, and Kenya.

**Figure 4:**
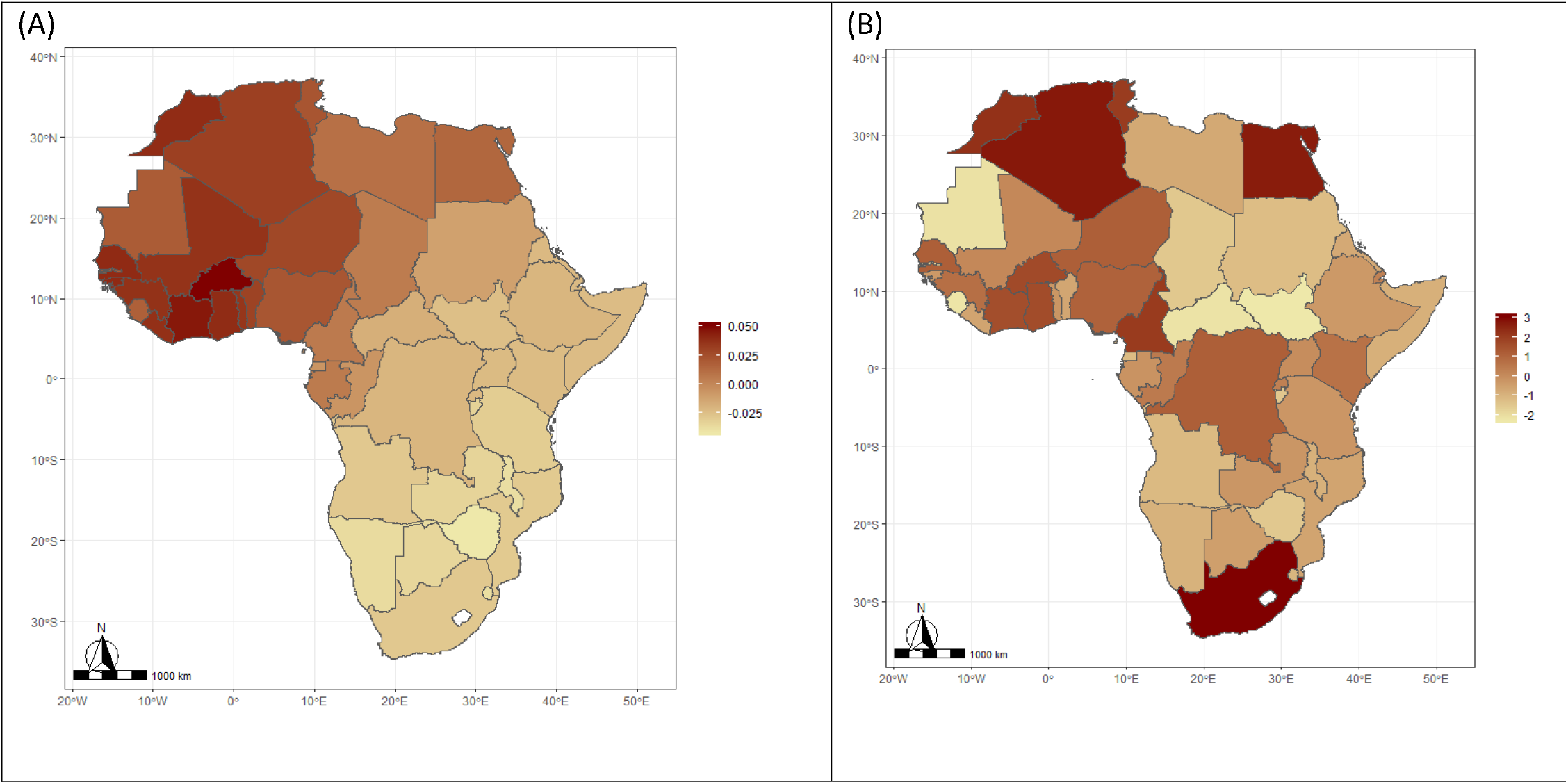
Structured (A) and unstructured (B) spatial effects for the mean of COVID-19 (lambda parameter) in Africa.

Results for the spatial patterns of the probability of no occurrence are presented in Figure 5 (A & B). The structured spatial effects show that neighboring countries in the southern and central Africa have the highest likelihood of not recording any cases of COVID-19. The map for the unstructured effect however reveals that the likelihood of not reporting a case was highest among Mauritania, Botswana, South Sudan, Burundi, Namibia, Libya, Chad, Central African Republic, Somalia, Malawi, Benin, Sierra Leone, The Gambia and Swaziland, but lowest for South Africa, Egypt, Algeria, Morocco, Tunisia, and Senegal. The temporal patterns presented in Figure 6 (A & B) displayed the posterior mean estimate (black) and 95% credible interval (blue). The Figure reveals that the frequency of occurrence has been on a consistent rise since the first case was reported up to around day 50, followed by a somewhat gradual decline for about three days after which there was evidence of another rise. On the other hand, the estimates for the likelihood of no occurrence decline sharply till day 50 and appear to flatten thereafter. Note that the wide credible intervals at the early days are evidence of few reported cases at those periods.

**Figure 5:**
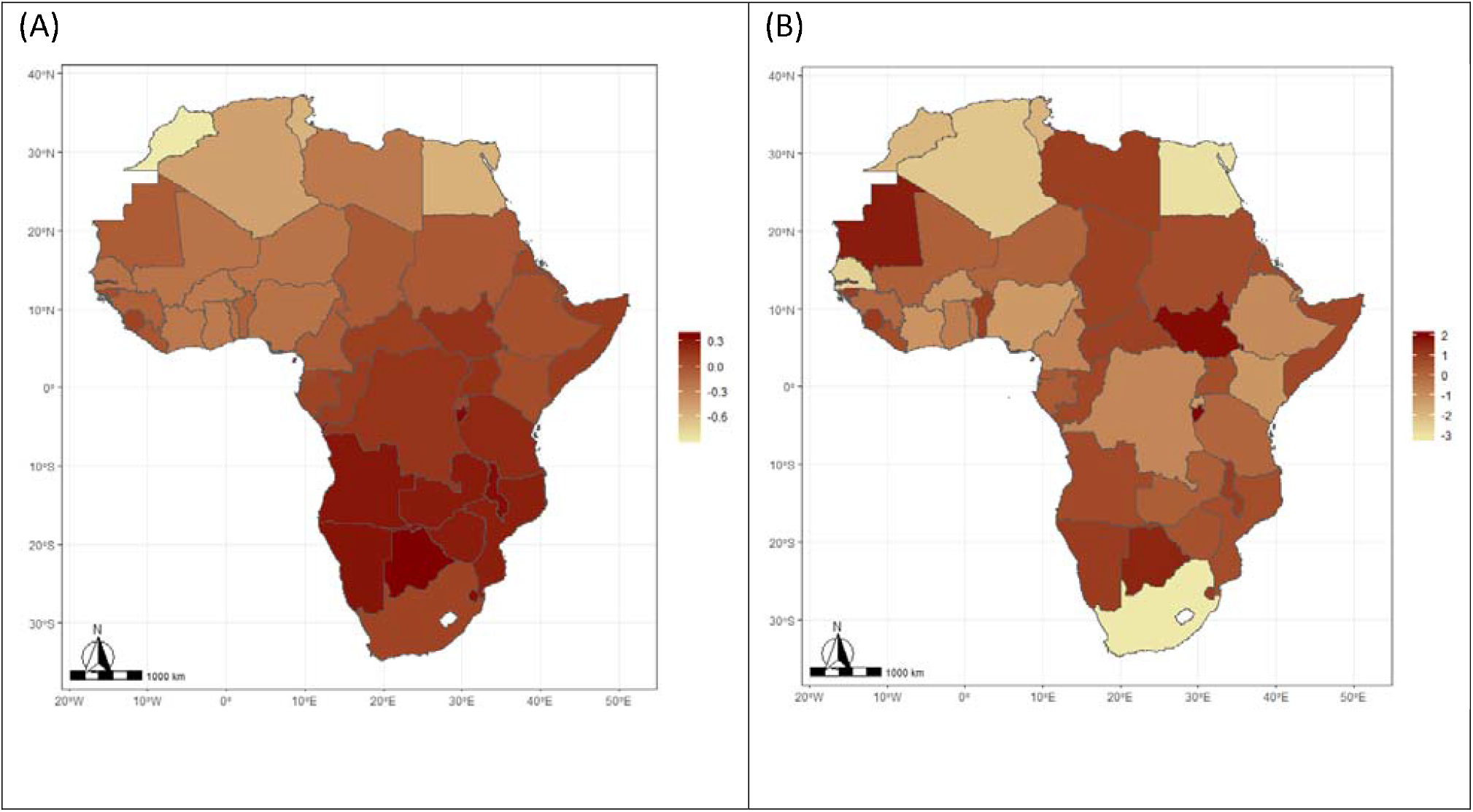
Structured (A) and unstructured (B) spatial effects for the probability of no occurrence of COVID-19 (pi parameter) in Africa.

**Figure 6:**
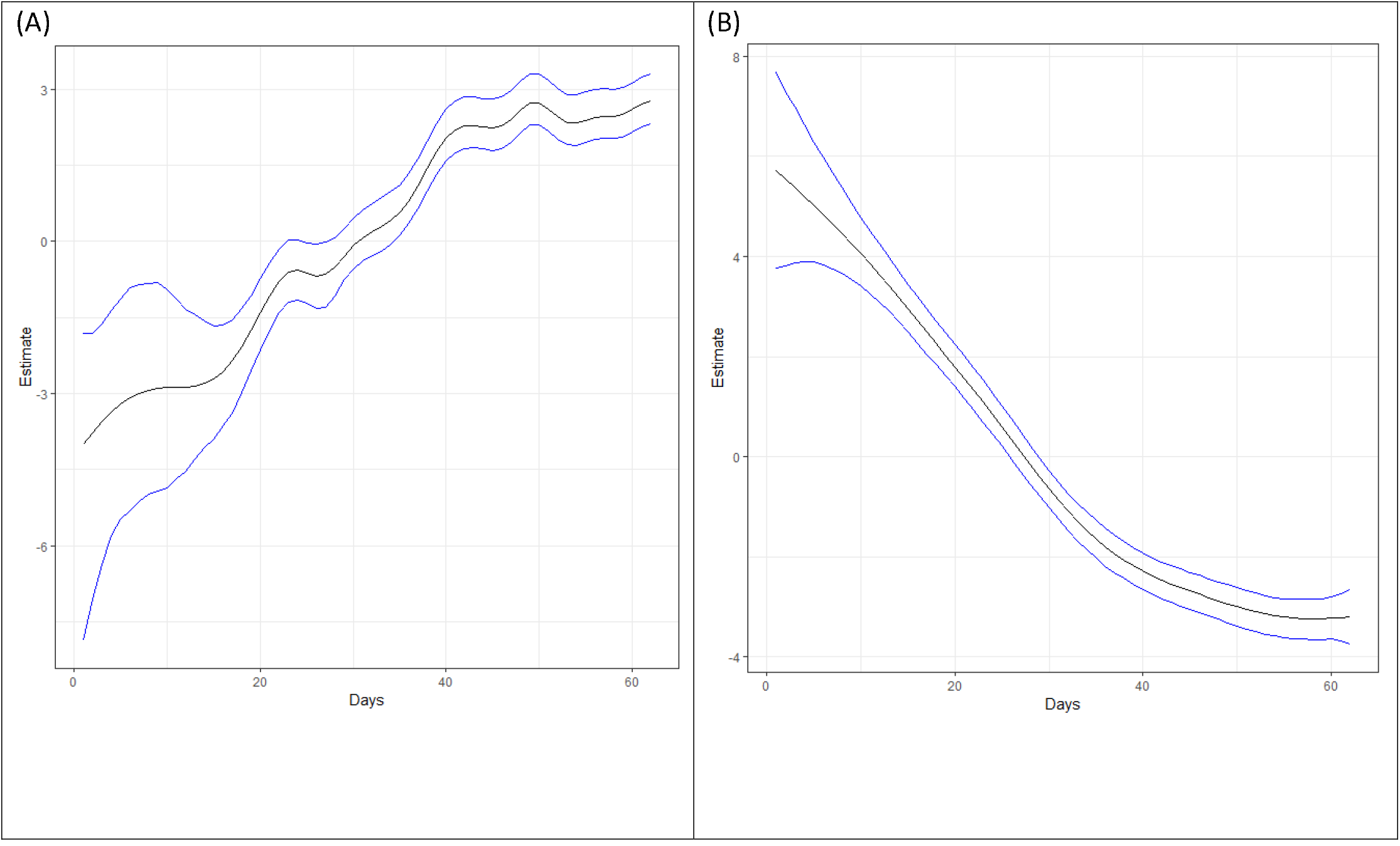
Temporal trend of COVID-19 for (A) mean number of occurrence and (B) likelihood of no occurrence.

## Discussion

This study has established that the burden of COVID-19 in Africa varies geographically with each country’s healthcare-related variables. As COVID-19 causes significant health and economic challenges globally, the impacts on Africa are still in their infancy. The present study reveals that the geographical spread of the disease in Africa and its relation to individual country health capacities. Findings from the spatio-temporal analysis reveal that the occurrence and burden of COVID-19 in Africa varied geographically with neighboring countries particularly in the western part of the continent which could imply that neighboring countries pose significant importation risk to each other. This is quite challenging but explains the reason why Africa countries should form a coalition to fight against COVID-19.

There are several possible reasons for the geographical distribution of COVID-19 across Africa. The first is the route of disease introduction to Africa. For example, when China was the only COVID-19 epicenter, the risk of COVID-19 importation from China to Africa was high for North Africa [6]. A study based on travel data from provinces in China, identified Egypt, Algeria, and South Africa as having the highest importation risk from China [5, 6]. However, as the epicenter changes from China to Italy and the US, the risk to other regions in Africa increases as there more African travelers from Europe and North America than Asia [5]. The second reason is the issue of border porosity in most African countries [19, 20]. The ease of people’s movement between borders could increase importations as seen in Nigeria where many returning Nigerians from Ivory Coast were diagnosed to have COVID-19. This is what happens with neighboring African countries as they endanger each other unless enhanced border and air-flight restrictions are put in place.

COVID-19 is a significant health issues because it can quickly overwhelm healthcare capacity if not checked. In this study, countries with more healthcare capacities measured by number of hospital beds and physicians have more cases. Healthcare capacity could use as a measure of a country’s wealth [21, 22], therefore, it is more likely that citizens of such countries have more tendencies for traveling overseas, thereby having a greater chance of importing COVID-19 and other infectious diseases on their return. Our finding is consistent with a previous study that suggested African countries with higher surveillance systems are more likely to identify a higher risk of disease importation [6]. This implies that additional public health capacity is needed for those countries that have limited resources to detect COVID-19 and undertake meaning contact tracing to curtail the rapid spread of the virus.

There have been warnings that some countries in Africa could be the next epicenters [23, 24]. Thus far, the burden of COVID-19 in Africa is low in comparison with Europe, Asia, and North America. There is a need for early introduction of interventions such as isolation, quarantine and social distancing [25]. However, many African countries are poor and whether these control measures will work as effectively as seen in China is still an open question [24].

## Conclusion

As the pandemic spreads, the African Centers for Disease Control have intensified investment in enhancing diagnostic and surveillance capacity across the countries [26]. Africa may only be able to fight this virus if conscientious efforts and support are garnered globally to battle COVID-19 [23]. We have shown the trajectory of COVID-19 in Africa is peaking up, with each African country posing a risk to its neighbors. The findings in this study will be useful in implementing epidemiological intervention based on heterogeneity of the disease patterns for allocation of resources and targeted intervention strategies

## Data Availability

we used publicly available daily number of confirmed COVID-19 cases reported by the World Health Organization (https://covid19.who.int)

## Supplementary materials

**Figure S1.**
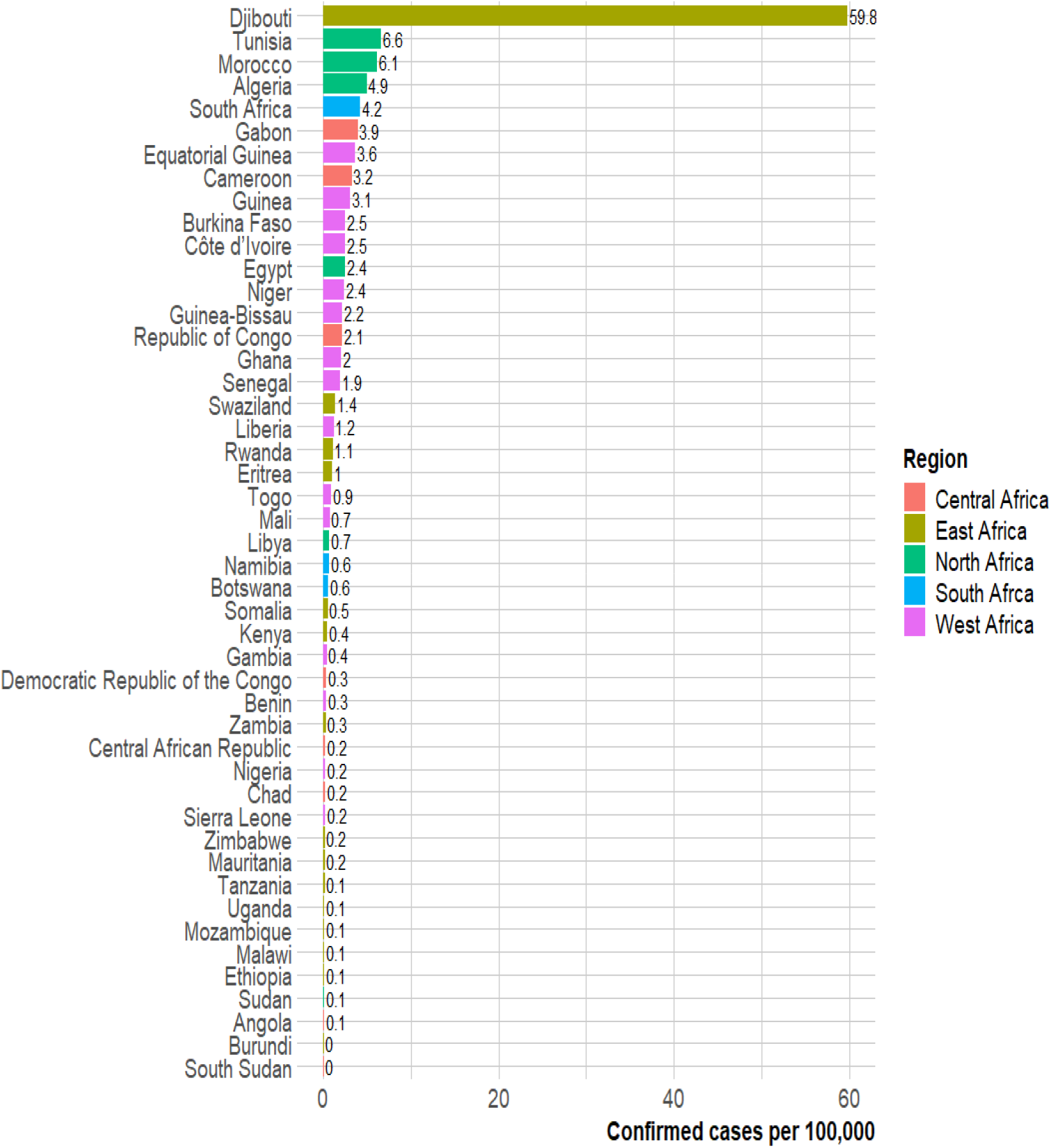
Burden (cases per 100,000 population) of COVID-19 across Africa as at 16^th^ April 2020.

## Notes

### Competing Interest Statement

The authors have declared no competing interest.

